# COVID-19 Vaccine Acceptance Among Health Care Workers in the United States

**DOI:** 10.1101/2021.01.03.21249184

**Authors:** Rahul Shekhar, Abu Baker Sheikh, Shubhra Upadhyay, Mriganka Singh, Saket Kottewar, Hamza Mir, Eileen Barrett, Suman Pal

## Abstract

**Background:** Acceptance of the COVID-19 vaccine will play a major role in combating the pandemic. Healthcare workers (HCWs) are amongst the first group to receive vaccination, so it is important to consider their attitudes about COVID-19 vaccination to better address barriers to widespread vaccination acceptance.

**Methods:** We conducted a cross sectional study to assess the attitude of HCWs toward COVID-19 vaccination. Data was collected between October 7th and November 9th, 2020. We received 4080 responses out of which 3479 were complete responses and were included in final analysis.

**Results:** 36% of respondents were willing to take the vaccine as soon as it became available while 56% were not sure or would wait to review more data. Vaccine acceptance increased with increasing age, education, and income level. Lower acceptance was noted in females (31%), Black (10%), Latinx (30%) and Conservative/Republican (21%) HCWs, and those working in a rural setting (26%). Direct medical care providers had higher vaccine acceptance (49%). Safety (69%), effectiveness (69%) and speed of development/approval (74%) were noted as the most common concerns regarding COVID-19 vaccination in our survey.

**Conclusion:** Immediate acceptance of a COVID-19 vaccine is low, with the majority of HCWs choosing to wait to review more data before deciding on personal vaccination. Overall attitudes toward vaccination were positive but specific concerns regarding COVID-19 vaccine are prevalent. Differences in vaccine acceptance were noted between individual and group characteristics which should be addressed to avoid exacerbating health inequities.

## Introduction

COVID-19 has rapidly become a major global public health crisis, affecting 86.4 million individuals, and causing 1.9 million deaths. The US has reported more than 21 million cases and 357,000 deaths. To curb this pandemic, apart from effective public health measures such as social distancing, wearing face masks, hand washing and avoidance of crowded indoor spaces, educating the general population, efficacious vaccination is emerging as essential to mitigating disease and death.^2-5^.

Uptake of any COVID-19 vaccine is an important challenge to address. In a recent survey, more than one-third of lay respondents were unsure or did not intend to take the vaccine^7^. Clinicians are an important source of information for vaccines and physician communication can improve adherence to vaccination recommendations ^8,9,10^. Thus, the role of healthcare workers (HCWs) becomes particularly important in advising patients and communities, and as well as through role modeling behavior. HCWs are prioritized among the high-risk groups who are considered as candidates for early vaccination. As such, it is important to consider HCW attitudes about COVID-19 vaccination to better address barriers to widespread vaccination.

## Methods

### Design

We conducted a cross sectional study to assess the attitude of HCWs toward COVID-19 vaccination. An online questionnaire was created using REDCap electronic data capture tools hosted at the University of New Mexico. The survey was modified from a previously published general population survey^7^ to capture more information pertinent to healthcare workers. The data was collected anonymously, and no personally identifying information was collected. This study was approved by University of New Mexico Hospitals Institutional Review Board.

### Sampling

A snowball sampling was utilized. The survey tool was distributed via links posted on social media platforms in various HCW groups and distributed to administrative leaders at five major hospital systems to disseminate among their employees. Data was collected between October 7th and November 9th, 2020.

### Participants

All adults (>18 years of age) working in a healthcare setting regardless of patient care contact and role in health care settings were eligible to participate in the study. Informed consent was obtained prior to enrollment in the study.

## Measures

Demographic information collected included age, gender, ethnicity, race, state of primary residence in majority of last six months, occupation, marital status, the number of household members excluding participant, annual household income, location of healthcare setting (rural, suburban or urban), education level and political orientation (Conservative/Republican, unaffiliated, Democrat/Liberal, or do not wish to answer).

Self-perceived risk of COVID-19 was gauged by the question “Do you think you are at risk of getting COVID-19 in the next 1 year?” The responses allowed for graded self-perceived risk (“No I am confident I won’t get infected”; “Yes I am concerned that I will get mild symptoms which will probably not require hospitalization”; “Yes I am concerned that I will get moderate symptoms which will probably need hospitalization”; “Yes I am concerned that I will get severe symptom which will probably require admission to the intensive care unit”; “I believe I already have the disease and I am immune to it (not diagnosed by a test)”; “No, I already have recovered and won’t get re-infected (diagnosed by a test)”.

Exposure to COVID-19 was assessed by the questions “Have you directly or indirectly taken care of the COVID-19 patients?” and “Have you, your family member or someone you know been diagnosed with COVID-19 (excluding your patients)?”

Acceptance of COVID-19 vaccine was assessed by the question “When COVID-19 vaccination becomes available, would you take it?” Participants could choose responses from among the options: “Yes, as soon as I can get it”; “Yes, only if it is required by employer”; “No, I will wait for 3 months to review safety profile”; “No, I will wait for 6 months to review safety profile”; “No, I will wait for at least 1 year to review safety profile”; “I will not get the vaccine”; “Not sure”.

Attitude toward vaccination was assessed by agreement with perception/concern statements. General perception about vaccines were assessed by statements “I do not believe vaccines work”; “I do not believe vaccines are safe”; “I do not get vaccinated for religious reasons”; “I do not get vaccinated for reasons of personal freedom/choice”; “I do not get vaccinated for a fear of needles/doctors/hospitals”. Concerns regarding COVID-19 vaccine were evaluated by statements “I am worried about the safety/adverse effects of COVID-19 vaccine”; “I am worried about effectiveness of COVID-19 vaccine”; “I am worried about the out of pocket cost/Insurance coverage of the vaccine”; “I am concerned about adverse effect of vaccine on my pre-existing conditions”; “I am worried about the rapidity of the development and approval of COVID-19 vaccine”; “I do not need the vaccine for my risk level”; “I am worried about the rapidity of the development and approval of COVID-19 vaccine”. Agreement was measured on a Likert scale from strongly disagree to strongly agree.

### Outcome

We divided HCWs into four major categories: Direct medical provider (DMP) which includes If the participant is a physician/resident/medical student/advanced practice provider (including Nurse Practitioner and physician’s assistant); Direct patient care provider (DPCP) including registered nurse /patient care technician/paramedic/rehabilitation services (respiratory, physical, occupational, or speech therapist), nutritionist, social worker, case manager, care coordinator, Administrative staff, and Others without direct patient care. If the participant is DMP their primary specialty was also recorded and is divided into primary medical, primary surgical diagnostic and others.

The primary outcome of the survey was whether HCWs are willing to take the COVID-19 vaccine or not. Responses were collected against willingness to “take it as soon as it becomes available” and “yes but only if it is required by employer” (grouped into one as yes responses), “wait for safety data review for 3 months”, “wait for safety data review for 6 months” or “wait for safety data review for a year” or “not sure” (grouped into one response “wait for review”), and “not willing” to take it.

We performed a multinomial logistic regression due to the overall description of the data. We split the sample into 3 groups according to the primary outcome variable, would a participant take the COVID-19 vaccination immediately, or would wait to review safety data, or would not take the vaccination at all. To analyze association between vaccine acceptance and participant characteristics, we used likelihood-ratio test leading to the derived chi-square and p-values. Statistical significance was assumed for p<0.05. We used the R programming language (R Foundation for Statistical Computing, Vienna, Austria. http://www.R-project.org/) to develop the model and analyze its results.

## Results

3,479 HCWs completed survey. Most participants were younger than 40 years of age (1877, 54%), female (2598, 75%), White (2803, 81%), completed a bachelor’s degree or higher (2788, 80%) identified as Democrat or Liberal (1521, 44%) and had no chronic medical conditions (2039, 59%). Most reported working in an urban area (2229, 64%), in primary medical and medical subspecialties (54%) and provided direct patient care (79%). The majority of participants perceived themselves to be at risk for acquiring COVID-19 (3043, 87%) and 21% think they will acquire serious disease requiring admission to hospital (747) but less than eight percent participant were confident that they would not get the disease (267, 8%). Roughly half of the HCWs have directly taken care of COVID-19 positive patients (1570, 45%). Most of the participants believed that COVID-19 vaccination should be voluntary (1665,48%). Additional sample characteristics are shown in Table 1.

**Table 1:** Participant characteristics.

|  |  |
| --- | --- |
| **Table 1: Participant characteristics** |  |
| **Variable** | **N = 3,479<sup>a</sup>** |
| **Age** |  |
| 18-30 years | 816<br>(23%) |
| 31-40 years | 1,061<br>(30%) |
| 41-50 years | 686<br>(20%) |
| 51-60 years | 571<br>(16%) |
| 61-70 years | 326<br>(9.4%) |
| > 70 years | 19 (0.5%) |
| **Gender** |  |
| Female | 2,598<br>(75%) |
| Male | 864<br>(25%) |
| Trans/ Gender non-binary/ not specified above | 7 (0.2%) |
| Do not wish to reply | 10 (0.3%) |
| **Ethnicity** |  |
| Hispanic or Latino | 560<br>(16%) |
| NOT Hispanic or Latino | 2,763<br>(79%) |
| Unknown / Not Reported | 36 (1.0%) |
| Do not wish to answer | 120<br>(3.4%) |
| **Race** |  |
| White or Caucasian | 2,803<br>(81%) |
| Asian | 218<br>(6.3%) |
| Black or African American | 74 (2.1%) |
| Native Americans/Alaska Native | 30 (0.9%) |
| Native Hawaiian or Other Pacific Islander | 6 (0.2%) |
| More Than One Race | 126<br>(3.6%) |
| Unknown / Other | 70 (2.0%) |
| Do not wish to answer | 152<br>(4.4%) |
| **State of Residence** |  |
| Midwest | 1,433<br>(41%) |
| North East | 81 (2.3%) |
| South | 314<br>(9.0%) |
| West | 1,651<br>(47%) |
| **Occupation** |  |
| Direct Patient Care Providers (DPCP) | 1,573<br>(45%) |
| Direct Medical Providers (DMP) | 1,207<br>(35%) |
| Administrative staff working in hospital without direct patient contact | 295<br>(8.5%) |
| Others without direct patient contact | 404<br>(12%) |
| **Primary Area of Work** |  |
| Primary medical and medical subspecialty | 1,882<br>(54%) |
| Primary surgical and surgical subspecialty | 363<br>(10%) |
| Diagnostic subspecialty | 246<br>(7.1%) |
| Others | 988<br>(28%) |
| **Annual Income** |  |
| < \$30,000 | 117<br>(3.4%) |
| \$30,001 - \$70,000 | 741<br>(21%) |
| \$70,001 - \$100,00 | 759<br>(22%) |
| \$100,001 - \$150,000 | 796<br>(23%) |
| >\$150,001 | 1,066<br>(31%) |
| **Health Care facility in** |  |
| Rural area | 293<br>(8.4%) |
| Suburban area | 957<br>(28%) |
| Urban area | 2,229<br>(64%) |
| **Education** |  |
| No formal education | 1 (<0.1%) |
| High school graduate, diploma or the equivalent (for example: GED) | 46<br>(1.3%) |
| Some college credit, no degree | 169<br>(4.9%) |
| Trade/technical/vocational training | 111<br>(3.2%) |
| Associate degree | 364<br>(10%) |
| Bachelor's degree | 1,046<br>(30%) |
| Master's degree | 606<br>(17%) |
| Professional degree | 297<br>(8.5%) |
| Doctorate degree | 839<br>(24%) |
| **Political Identification** |  |
| Conservative-Republican | 746<br>(21%) |
| Democrat-Liberal | 1,519<br>(44%) |
| Unaffiliated | 640<br>(18%) |
| Do not wish to answer | 574<br>(16%) |
| **Medical comorbidities** |  |
| No Chronic Condition | 2,039<br>(59%) |
| Diabetes Mellitus Type-1/Type-2 | 136<br>(3.9%) |
| Heart Disease | 36 (1.0%) |
| Hypertension | 449<br>(13%) |
| COPD Asthma - Lung Disease | 318<br>(9.1%) |
| Obesity BMI>30 | 461<br>(13%) |
| Cancer | 125<br>(3.6%) |
| Immuno-compromised/on immunosuppressants | 110<br>(3.2%) |
| Smoking | 288<br>(8.3%) |
| Other Medical Conditions | 505<br>(15%) |
| **Have you, your family member or someone you know been diagnosed with COVID-19 (Excluding your patients)** |  |
| I was Diagnosed with COVID-19 | 90 (2.6%) |
| Family Member was Diagnosed with COVID-19 | 447<br>(13%) |
| Someone Personal was Diagnosed with COVID-19 | 1,793<br>(52%) |
| No one Personal was Diagnosed with COVID-19 | 1,400<br>(40%) |
| **Do you think you are at risk of getting COVID-19 in the next 1 year?** |  |
| I believe I already have the disease and I am immune to it (Not diagnosed by a test) | 138<br>(4.0%) |
| No, I am confident I won't get infected | 266<br>(7.6%) |
| No, I already have recovered and won't get re-infected (Diagnosed by a test) | 34 (1.0%) |
| Yes, I am concerned that I will get mild symptoms which will probably not require hospitalization | 2,294<br>(66%) |
| Yes, I am concerned that I will get moderate symptoms which will probably need hospitalization | 572<br>(16%) |
| Yes, I am concerned that I will get severe symptom which will probably require admission to the Intensive care unit | 175<br>(5.0%) |
| **Have you directly or indirectly taken care of the COVID-19 patients?** |  |
| No | 1,263<br>(36%) |
| Yes, but no direct patient contact | 646<br>(19%) |
| Yes, I have direct patient contact | 1,570<br>(45%) |
| **Would you take the COVID-19 Vaccine** |  |
| No | 279<br>(8.0%) |
| Wait for Review | 1,953<br>(56%) |
| Yes | 1,247<br>(36%) |
| **Would you advise friends and family to get vaccinated for COVID-19?** |  |
| No | 519<br>(15%) |
| Not sure | 1,376<br>(40%) |
| Yes | 1,584<br>(46%) |
| **COVID-19 Vaccine for health care workers should be:** |  |
| Mandated by the employer, like Influenza vaccine | 792<br>(23%) |
| Mandated by the Federal government for all health care workers | 338<br>(9.7%) |
| Mandated by the State government for all health care workers | 99 (2.8%) |
| Not sure | 585<br>(17%) |
| Voluntary | 1,665<br>(48%) |
| Statistics presented: n (%) |  |
| Direct Patient Care Provider: Registered Nurse / Patient care technician/ Paramedics/ Rehab services – Respiratory therapist/Physical therapist/Occupation therapist/Speech/ Nutritionist/ social workers/ care coordinators |  |
| Direct Medical Provider: Physician/<br>Resident/Medical students and Advanced practice provider (Nurse Practitioner<br>/ Physician's assistant |  |
| Diagnostic Subspeciality: Radiology / Cath/ Endoscopy/ Pulmonary/ Echo/ Sleep Laboratory Technicians |  |

Only about one-third (1247, 36%) of respondents were willing to take a COVID-19 vaccine as soon as it became available at the time of the survey. A majority of the HCW were not sure or would wait to review safety data before getting vaccinated (1953, 56%). Among the respondents who want to wait, 11% will like to wait for 3 months, 10% will like to wait for 6 months and 20% would like to wait at least 1 year. Only 8% (279) of respondents were unwilling to take the vaccine at all.

A significant association was noted between the choice that participants make about receiving COVID-19 vaccination and multiple predictor variables (Table 2). We note that acceptance of COVID-19 vaccination increased with increasing age. In the 18-30 age group only 34% of respondents were willing to take COVID-19 vaccine as soon as it became available which increased to 47% in the >70 age group. A similar trend was noted with education and income level; increasing education and income levels represent a higher proportion of HCWs willing to take the vaccine as soon as it becomes available.

**Table 2:** Participant characteristics by vaccine acceptance.

| Variable | No, N =<br>279<sup>1</sup> | Wait for<br>Review, N =<br>1,953<sup>1</sup> | Yes, N =<br>1,247<sup>1</sup> | p-value<sup>2</sup> |
| --- | --- | --- | --- | --- |
| **Age** |  |  |  | <0.001* |
| 18-30 years | 72 (8.8%) | 467 (57%) | 277<br>(34%) |  |
| 31-40 years | 83 (7.8%) | 615 (58%) | 363<br>(34%) |  |
| 41-50 years | 70 (10%) | 389 (57%) | 227<br>(33%) |  |
| 51-60 years | 43 (7.5%) | 306 (54%) | 222<br>(39%) |  |
| 61-70 years | 10 (3.1%) | 167 (51%) | 149<br>(46%) |  |
| > 70 years | 1 (5.3%) | 9 (47%) | 9 (47%) |  |
| **Gender** |  |  |  | <0.001* |
| Female | 240<br>(9.2%) | 1,540 (59%) | 818<br>(31%) |  |
| Male | 37 (4.3%) | 402 (47%) | 425 (49%) |  |
| Trans/ Gender non-binary/ not specified above | 0 (0%) | 4 (57%) | 3 (43%) |  |
| Do not wish to reply | 2 (20%) | 7 (70%) | 1 (10%) |  |
| **Ethnicity** |  |  |  | <0.001* |
| Hispanic or Latino | 55 (9.8%) | 337 (60%) | 168 (30%) |  |
| NOT Hispanic or Latino | 191 (6.9%) | 1,536 (56%) | 1,036 (37%) |  |
| Unknown / Not Reported | 8 (22%) | 16 (44%) | 12 (33%) |  |
| Do not wish to answer | 25 (21%) | 64 (53%) | 31 (26%) |  |
| **Race** |  |  |  | <0.001* |
| White or Caucasian | 221 (7.9%) | 1,545 (55%) | 1,037 (37%) |  |
| Asian | 1 (0.5%) | 122 (56%) | 95 (44%) |  |
| Black or African American | 12 (16%) | 48 (65%) | 14 (19%) |  |
| Native Americans/Alaska Native | 3 (10%) | 24 (80%) | 3 (10%) |  |
| Native Hawaiian or Other Pacific Islander | 0 (0%) | 6 (100%) | 0 (0%) |  |
| More Than One Race | 4 (3.2%) | 82 (65%) | 40 (32%) |  |
| Unknown / Other | 9 (13%) | 41 (59%) | 20 (29%) |  |
| Do not wish to answer | 29 (19%) | 85 (56%) | 38 (25%) |  |
| **State of Residence** |  |  |  | 0.003* |
| Midwest | 146<br>(10%) | 776 (54%) | 511<br>(36%) |  |
| North East | 2 (2.5%) | 45 (56%) | 34 (42%) |  |
| South | 11 (3.5%) | 151 (48%) | 152<br>(48%) |  |
| West | 120<br>(7.3%) | 981 (59%) | 550<br>(33%) |  |
| **Occupation** |  |  |  | <0.001* |
| Direct patient care providers (DPCP) | 187<br>(12%) | 969 (62%) | 417<br>(27%) |  |
| Direct Medical Provider (DMP) | 30 (2.5%) | 582 (48%) | 595<br>(49%) |  |
| Administrative staff working in hospital without direct patient contact | 25 (8.5%) | 170 (58%) | 100<br>(34%) |  |
| Others without direct patient contact | 37 (9.2%) | 232 (57%) | 135<br>(33%) |  |
| **Annual Income** |  |  |  | <0.001* |
| < \$30,000 | 19 (16%) | 53 (45%) | 45 (38%) | |
| \$30,001 - \$70,000 | 81 (11%) | 420 (57%) | 240<br>(32%) | |
| \$70,001 - \$100,00 | 80 (11%) | 463 (61%) | 216 (28%) | |
| \$100,001 - \$150,000 | 60 (7.5%) | 472 (59%) | 264 (33%) | |
| >\$150,001 | 39 (3.7%) | 545 (51%) | 482 (45%) | |
| **Health Care facility in** |  |  |  | <0.001* |
| Rural area | 52 (18%) | 164 (56%) | 77 (26%) |  |
| Suburban area | 98 (10%) | 523 (55%) | 336 (35%) |  |
| Urban area | 129 (5.8%) | 1,266 (57%) | 834 (37%) |  |
| **Education** |  |  |  | <0.001* |
| No formal education | 0 (0%) | 1 (100%) | 0 (0%) |  |
| High school graduate, diploma, or the equivalent (for example: GED) | 9 (20%) | 26 (57%) | 11 (24%) |  |
| Some college credit, no degree | 24 (14%) | 94 (56%) | 51 (30%) |  |
| Trade/technical/vocational training | 18 (16%) | 69 (62%) | 24 (22%) |  |
| Associate degree | 56 (15%) | 220 (60%) | 88 (24%) |  |
| Bachelor's degree | 109 (10%) | 627 (60%) | 310 (30%) |  |
| Master's degree | 40 (6.6%) | 362 (60%) | 204 (34%) |  |
| Professional degree | 6 (2.0%) | 139 (47%) | 152 (51%) |  |
| Doctorate degree | 17 (2.0%) | 415 (49%) | 407 (49%) |  |
| **Political Identification** |  |  |  | <0.001* |
| Conservative-Republican | 93 (12%) | 390 (52%) | 263 (35%) |  |
| Democrat-Liberal | 42 (2.8%) | 833 (55%) | 644 (42%) |  |
| Unaffiliated | 48 (7.5%) | 373 (58%) | 219 (34%) |  |
| Do not wish to answer | 96 (17%) | 357 (62%) | 121 (21%) |  |
| **Did you get the Influenza vaccine Last Year?** |  |  |  | <0.001* |
| No | 50 (43%) | 56 (48%) | 10 (8.6%) |  |
| Yes | 229 (6.8%) | 1,897 (56%) | 1,237 (37%) |  |
| **If you have Children under 18 years old, Have they been vaccinated for other diseases** |  |  |  | <0.001* |
| No | 38 (26%) | 67 (46%) | 41 (28%) |  |
| Not Applicable | 120 (6.0%) | 1,132 (56%) | 754 (38%) |  |
| Yes | 121 (9.1%) | 754 (57%) | 452 (34%) |  |
| **Medical Condition = Chronic Condition** |  |  |  | 0.046 |
| No | 107 (7.4%) | 819 (57%) | 514 (36%) |  |
| Yes | 172<br>(8.4%) | 1,134 (56%) | 733<br>(36%) |  |
| **Diabetes Mellitus Type-1/Type-2** |  |  |  | 0.076 |
| No | 272<br>(8.1%) | 1,876 (56%) | 1,195<br>(36%) |  |
| Yes | 7 (5.1%) | 77 (57%) | 52 (38%) |  |
| **Heart Disease** |  |  |  | 0.067 |
| No | 276<br>(8.0%) | 1,935 (56%) | 1,232<br>(36%) |  |
| Yes | 3 (8.3%) | 18 (50%) | 15 (42%) |  |
| **Hypertension** |  |  |  | 0.8 |
| No | 253<br>(8.3%) | 1,701 (56%) | 1,076<br>(36%) |  |
| Yes | 26 (5.8%) | 252 (56%) | 171<br>(38%) |  |
| **COPD Asthma - Lung Disease** |  |  |  | 0.701 |
| No | 258<br>(8.2%) | 1,778 (56%) | 1,125<br>(36%) |  |
| Yes | 21 (6.6%) | 175 (55%) | 122<br>(38%) |  |
| **Obesity BMI>30** |  |  |  | 0.022* |
| No | 250<br>(8.3%) | 1,671 (55%) | 1,097<br>(36%) |  |
| Yes | 29 (6.3%) | 282 (61%) | 150<br>(33%) |  |
| **Cancer** |  |  |  | 0.399 |
| No | 274<br>(8.2%) | 1,882 (56%) | 1,198<br>(36%) |  |
| Yes | 5 (4.0%) | 71 (57%) | 49 (39%) |  |
| **Immuno-compromised/on immunosuppressants** |  |  |  | 0.354 |
| No | 269<br>(8.0%) | 1,889 (56%) | 1,211<br>(36%) |  |
| Yes | 10 (9.1%) | 64 (58%) | 36 (33%) |  |
| **Smoking** |  |  |  | 0.013* |
| No | 262<br>(8.2%) | 1,778 (56%) | 1,151<br>(36%) |  |
| Yes | 17 (5.9%) | 175 (61%) | 96 (33%) |  |
| **Other Medical Conditions** |  |  |  | 0.629 |
| No | 227<br>(7.6%) | 1,663 (56%) | 1,084<br>(36%) |  |
| Yes | 52 (10%) | 290 (57%) | 163<br>(32%) |  |
| **I was Diagnosed with COVID-19** |  |  |  | 0.009* |
| No | 272<br>(8.0%) | 1,901 (56%) | 1,216<br>(36%) |  |
| Yes | 7 (7.8%) | 52 (58%) | 31 (34%) |  |
| **Family Member was Diagnosed with COVID-19** |  |  |  | 0.695 |
| No | 244<br>(8.0%) | 1,712 (56%) | 1,076<br>(35%) |  |
| Yes | 35 (7.8%) | 241 (54%) | 171<br>(38%) |  |
| **Someone Personal was Diagnosed with COVID-19** |  |  |  | 0.003* |
| No | 168<br>(10.0%) | 924 (55%) | 594<br>(35%) |  |
| Yes | 111<br>(6.2%) | 1,029 (57%) | 653<br>(36%) |  |
| **No one Personal was Diagnosed with COVID-19** |  |  |  | 0.143 |
| No | 134<br>(6.4%) | 1,181 (57%) | 764<br>(37%) |  |
| Yes | 145<br>(10%) | 772 (55%) | 483<br>(34%) |  |
| **Do you think you are at risk of getting COVID-19 in next 1 year?** |  |  |  | <0.001* |
| I believe I already have the disease and I am immune to it<br>(Not diagnosed by a test) | 30 (22%) | 68 (49%) | 40 (29%) |  |
| No, I am confident I won't get infected | 71 (27%) | 128 (48%) | 67 (25%) |  |
| No, I already have recovered and won't get re-infected<br>(Diagnosed by a test) | 4 (12%) | 18 (53%) | 12 (35%) |  |
| Yes, I am concerned that I will get mild symptoms which will probably not require hospitalization | 153<br>(6.7%) | 1,283 (56%) | 858<br>(37%) |  |
| Yes, I am concerned that I will get moderate symptoms which will probably need hospitalization | 15 (2.6%) | 350 (61%) | 207<br>(36%) |  |
| Yes, I am concerned that I will get severe symptom which will probably require admission to the Intensive care unit | 6 (3.4%) | 106 (61%) | 63 (36%) |  |
| **Have you directly or indirectly taken care of the COVID-19 patients?** |  |  |  | <0.001* |
| No | 116<br>(9.2%) | 714 (57%) | 433<br>(34%) |  |
| Yes, but no direct patient contact | 45 (7.0%) | 358 (55%) | 243<br>(38%) |  |
| Yes, I have direct patient contact | 118<br>(7.5%) | 881 (56%) | 571<br>(36%) |  |
| **I would get the vaccine to prevent COVID-19 in myself** |  |  |  | <0.001* |
| No | 277<br>(20%) | 816 (58%) | 324<br>(23%) |  |
| Yes | 2<br>(<0.1%) | 1,137 (55%) | 923<br>(45%) |  |
| **I would get the vaccine to prevent COVID-19 in friends and family members** |  |  |  | <0.001* |
| No | 276<br>(21%) | 723 (56%) | 286<br>(22%) |  |
| Yes | 3 (0.1%) | 1,230 (56%) | 961<br>(44%) |  |
| **I would get the vaccine to prevent COVID-19 in community** |  |  |  | <0.001* |
| No | 277<br>(26%) | 560 (52%) | 237<br>(22%) |  |
| Yes | 2<br>(<0.1%) | 1,393 (58%) | 1,010<br>(42%) |  |
| **I would not get the vaccine** |  |  |  | <0.001* |
| No | 3<br>(<0.1%) | 1,787 (59%) | 1,235<br>(41%) |  |
| Yes | 276<br>(61%) | 166 (37%) | 12 (2.6%) |  |
| **Would you advise friends and family to get vaccinated for COVID-19?** |  |  |  | <0.001* |
| No | 230<br>(44%) | 272 (52%) | 17 (3.3%) |  |
| Not sure | 49 (3.6%) | 1,162 (84%) | 165<br>(12%) |  |
| Yes | 0 (0%) | 519 (33%) | 1,065<br>(67%) |  |
| **COVID-19 Vaccine for health care workers should be:** |  |  |  | <0.001* |
| Mandated by the employer, like Influenza vaccine | 0 (0%) | 245 (31%) | 547<br>(69%) |  |
| Mandated by the Federal government for all health care workers | 0 (0%) | 90 (27%) | 248<br>(73%) |  |
| Mandated by the State government for all health care workers | 0 (0%) | 33 (33%) | 66 (67%) |  |
| Not sure | 5 (0.9%) | 457 (78%) | 123 (21%) |  |
| Voluntary | 274 (16%) | 1,128 (68%) | 263 (16%) |  |
| Statistics presented: n (%) |  |  |  |  |
| Statistical tests performed: Likelihood-Ratio Test, Chi-Square Test |  |  |  |  |
| Direct Patient Care Provider: Registered Nurse / Patient care technician/ Paramedics/ Rehab services – Respiratory therapist/Physical therapist/Occupation therapist/Speech/ Nutritionist/ social workers/ care coordinators |  |  |  |  |
| Direct Medical Provider: Physician/ Resident/Medical students and Advanced practice provider (Nurse Practitioner / Physician's assistant) |  |  |  |  |
| Diagnostic Subspecialty: Radiology / Cath/ Endoscopy/ Pulmonary/ Echo/ Sleep Laboratory Technicians |  |  |  |  |

Differences in vaccine acceptance were noted by gender and racial identity. Female HCWs had lower vaccine acceptance at 31% compared to male HCWs (49%) and trans/non-binary HCWs (43%). Black HCWs had lower acceptance (19%) with the majority choosing to wait to review safety data (65%) whereas Asian HCWs had high vaccine acceptance (44%). A majority of Native American HCWs (80%) and all Native Hawaiian/Other pacific islander HCWs (100%) chose to wait to review data for COVID vaccine. Vaccine acceptance was lower among those identifying as Hispanic or Latino (30%). Geographical variation in vaccine acceptance was also noted in our survey with West having the lowest (33%) and South having the highest (48%) vaccine acceptance. HCWs employed in rural settings had lower acceptance (26%) of the vaccine. Those identifying as a Democrat/Liberal had higher vaccine acceptance (42%). HCWs who believe themselves to be immune to COVID-19 and those who feel confident they will not get infected had the highest rates of refusing COVID-19 vaccination at 22% and 27% respectively. HCWs who had not taken care of COVID-19 patients had higher rates of vaccine refusal (9.2%).

Vaccine acceptance varied by occupational role in healthcare. DMPs had higher vaccine acceptance (49%) than administrative staff (34%) and others without direct patient care (33%). DPCPs had the lowest vaccine acceptance (27%) with nearly two-thirds (62%) of DPCP choosing to wait to review safety data.

Acceptance of COVID-19 vaccine was also associated with a plan to recommend vaccination to friends and family, and with a higher likelihood of wanting COVID-19 vaccine for HCWs to be mandatory. Of note, a large majority of respondents who plan not to get a COVID-19 vaccine would also not recommend the vaccine to friends or family and want vaccination to be voluntary.

While overall concerns regarding vaccination in general were low, concerns regarding COVID-19 vaccines were prevalent (Figure 1). Most HCWs believe that in general vaccination works (90%), is safe (86%) and did not mention personal (87%) or religious belief (95%) as a reason for not vaccinating. Most participants endorsed concerns (agree or strongly agree) about vaccine safety/adverse effects (69%), effectiveness (69%), and rapidity of development/approval (74%). A majority of HCWs trust their doctors and healthcare professionals recommending the COVID-19 vaccine (73%) but nearly half of the respondents do not trust information provided by the government about COVID-19 and its severity (46%) and one-third do not trust regulatory authorities like CDC or FDA overseeing the vaccine development and safety (34%).

**Fig. 1.**
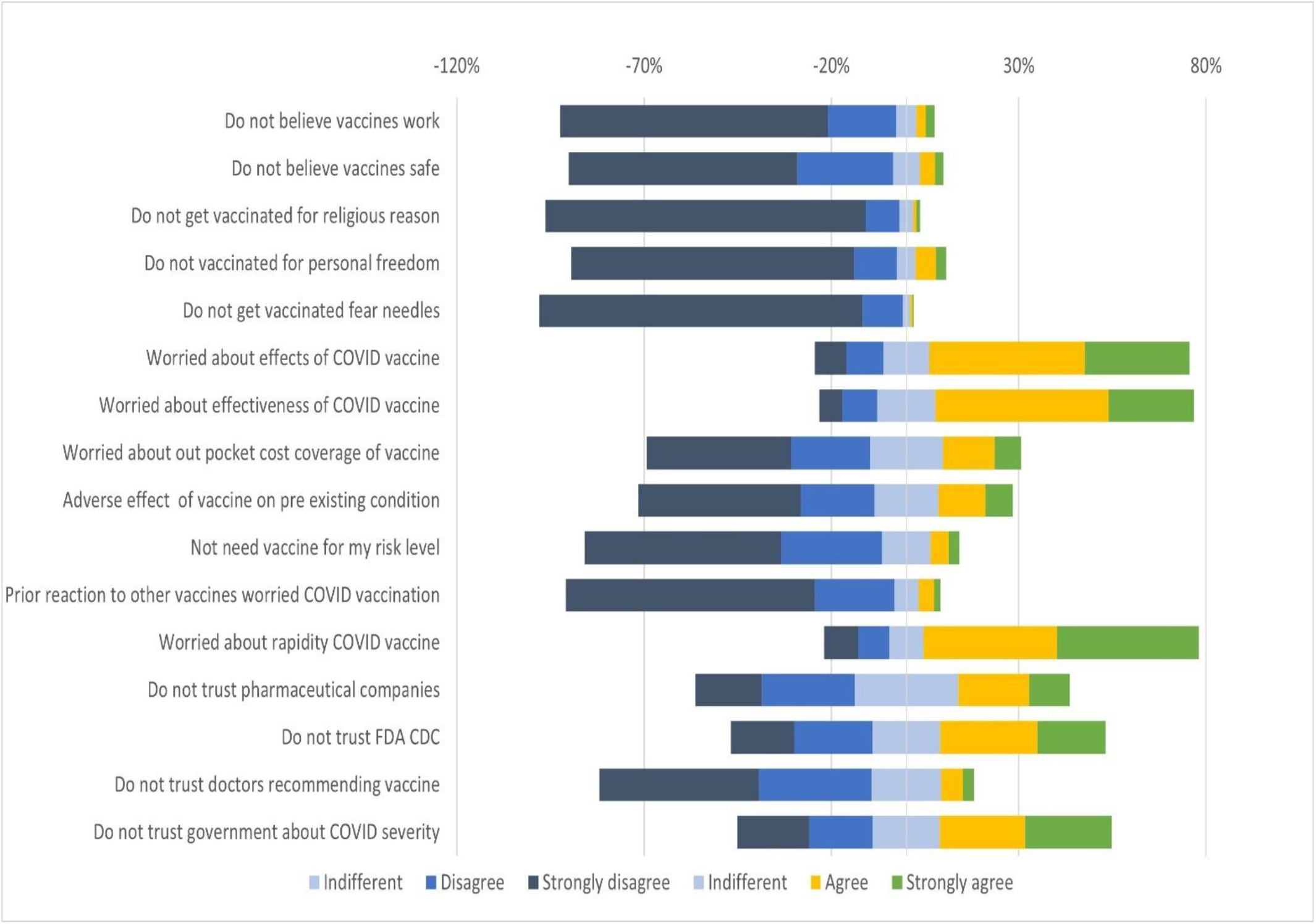

## Discussion

Since the announcement of efforts to develop a COVID-19 vaccine, several surveys have been conducted to gauge public perception and acceptance of the vaccine through 2020. Most surveys have focused on the general population. However, the rollout of the vaccine is tiered to various subgroups of the population based on limited availability, and HCWs are among the first subgroups of the US population to have access to the vaccine. HCWs are also likely to be an important source of information about the vaccine for the general population. As such, it is crucial to assess predictors of vaccine acceptance among HCWs which will help institutions and policy makers target resources to maximize uptake. To the best of our knowledge, this is the largest survey of HCWs in the US about COVID-19 vaccination.

In our survey, only one-third of respondents were amenable to COVID-19 vaccination immediately, while more than half of respondents preferred to defer their decision until reviewing more data. This contrasts with the findings of a general population survey in April 2020^7^ that reported half the participants were willing to take the vaccine. Another study in May 2020 reported 67% acceptance of COVID-19 vaccination among US adults^11^. The high percentage of respondents waiting to review more data in our study is expected as HCWs are more likely to base healthcare decisions on published scientific literature of efficacy and safety which was underway during the time of our survey. This also highlights the importance of publication and dissemination of scientific data regarding the vaccine which will be a crucial factor to determine eventual uptake of the vaccine among HCWs.

Overall, only 8% respondents said they would refuse the COVID-19 vaccine which shows a potential for high uptake of the vaccine among HCWs. Increasing vaccination acceptance has substantial benefits. To eventually slow down the spread of COVID-19 and its mortality, it is imperative to achieve herd immunity by vaccination before immunity by natural infection. With Ro of 3, the threshold herd immunity will be achieved by immunizing at least 70% of the population assuming the vaccine is 100% effective ^(17)^. With recent Emergency Use Authorizations by the Food and Drug Administration of 2 vaccines in the US reported to have almost 95% effectiveness in phase III clinical trials, this number could be even higher. Thus, it is imperative to vaccinate a maximum number of HCWs to prevent infection among HCWs and loss of critical workforce. Studies have shown that vaccination of HCWs with influenza vaccine decreases mortality and absenteeism^19-22^. It would be reasonable to expect a similar benefit with COVID-19 vaccination. It is important to note that 97% of HCWs in our survey had received an influenza vaccine in the previous year indicating a generally favorable perception of vaccination.

The low initial acceptance of COVID-19 vaccine among HCWs could also have broader consequences. Studies have shown that HCWs who are vaccinated are more likely to recommend vaccines to friends, family, and their patients^12-14^. This has also been borne out in our study where we see a strong association among HCWs who plan to be vaccinated and plan to recommend the vaccine to friends and family.

COVID-19 vaccine acceptance increased with increasing age, income, and education level. Black race also had lower vaccine acceptance while Asian race had higher vaccine acceptance. Vaccine acceptance was also lower in HCWs identifying as Hispanic or Latino. This mirrors the trends seen with general population surveys ^7,11^. The toll of COVID-19 pandemic has disproportionately affected low-income communities and Black, Indigenous, and people of color. ^(18)^. Lower vaccine uptake could exacerbate the health inequities among these communities. Targeted messaging and outreach would be required to achieve higher vaccination rates. It will also be important to understand and address the factors driving low vaccine acceptance which would be instrumental in addressing key concerns and directing resources to increase uptake.

HCWs identifying as Conservative or Republican had lower acceptance of the vaccine whereas those identifying as Liberal or Democrat had higher vaccine acceptance. This is in line with other general US population surveys^7,15,16.^ This difference could be in part due to differential messaging regarding vaccines in news and social media targeting these populations and in part due to very divergent responses to the pandemic by political leaders of major parties ^23^. A unified and consistent messaging from political leaders will be essential to bridge this gap.

Vaccine acceptance was higher among HCWs involved in direct patient care and in HCWs with chronic medical conditions. This could mirror trends in perceived risk of COVID-19 infection. More HCWs who perceived themselves to be at risk for COVID-19 infection were willing to accept than refuse the vaccine. However, even among HCWs directly involved in patient care, vaccine acceptance was also lower among HCWs identified as DPCP than among DMP. This is concerning since DPCP (such as nurses, respiratory therapists, etc.) often have more direct and prolonged patient contact. They are, therefore, at high risk of infection. DPCP are also one of the key resources of the healthcare system where critical shortages have been noted during the pandemic. They therefore represent a key subgroup whose health is essential to continue the care of patients with COVID-19 and understanding and addressing their concerns will be crucial.

To understand the factors driving vaccine uptake, we assessed HCWs attitude toward vaccination and toward COVID-19 vaccine. Concerns regarding vaccination in general were low in our study, consistent with other studies that show generally positive attitudes of healthcare workers toward vaccination. However, concerns specific to COVID-19 vaccination were prevalent. We found frequent concerns regarding vaccine efficacy, adverse effects, and rapidity of development. This was particularly noted among HCWs who do not plan to take the COVID-19 vaccine. Of note, while HCWs who did not want to be vaccinated reported poor trust in regulatory authorities and government, their trust in medical professionals prescribing the vaccine was somewhat higher. This could suggest an important role for dissemination of information through medical agencies and professional societies to increase uptake among HCWs.

A major strength of our study is the large sample of HCWs surveyed. Our survey population is also diverse with representation from different genders, age groups, ethnic and racial backgrounds, and roles in healthcare.

We recognize limitations of our study. Due to sampling methods, our study population may not be representative of all US HCWs. Social desirability bias may also affect the interpretation of our study results, though the responses were anonymized to minimize this. Most importantly, our study was conducted when information regarding COVID-19 vaccines under development were limited and findings of the clinical trials had not been made public. As such, it is possible that with this information now being publicly available, both vaccine acceptance and attitude toward COVID-19 vaccines have changed.

## Data Availability

The study participant data will be stored for six years at UNMH red cap for 6 years after the publication.

## References

1. COVID-19 Map - Johns Hopkins Coronavirus Resource Center. Johns Hopkins Coronavirus Resource Center. https://coronavirus.jhu.edu/map.html. Published 2020. Accessed January 5, 2021.

2. Adaptive Phase IB-II Randomized Clinical Trial Of Preventive Vaccine Consisting Of Autologous Dendritic Cells Loaded With Antigens From Severe Acute Respiratory Syndrome Coronavirus-2 (SARS-CoV-2), With Or Without GM-CSF, In Subjects Negative For COVID-19 Infection And Anti-SARS-CoV-2 Antibodies. ClinicalTrials.gov identifier (NCT number): NCT04386252. https://clinicaltrials.gov/ct2/show/NCT04386252

3. A Randomized, Double-blind, Placebo-controlled Phase 3 Study to Assess the Efficacy and Safety of Ad26.COV2.S for the Prevention of SARS-CoV-2-mediated COVID-19 in Adults Aged 18 Years and Older. ClinicalTrials.gov Identifier: NCT04505722. https://clinicaltrials.gov/ct2/show/NCT04505722

4. A Phase 2a, Randomized, Observer-Blind, Placebo Controlled, Dose-Confirmation Study to Evaluate the Safety, Reactogenicity, and Immunogenicity of mRNA-1273 SARS-COV-2 Vaccine in Adults Aged 18 Years and Older. ClinicalTrials.gov Identifier: NCT04405076. https://clinicaltrials.gov/ct2/show/NCT04405076

5. A Phase 1/Phase 2, Randomized, Double-Blind, Placebo-Controlled, Dose-Ranging Trial to Evaluate the Safety, Tolerability and Immunogenicity of V591 (COVID-19 Vaccine) in Healthy Younger and Older Participants. ClinicalTrials.gov Identifier: NCT04498247. https://www.clinicaltrials.gov/ct2/show/NCT04498247

6. Phadke VK, Bednarczyk RA, Salmon DA, Omer SB. Association Between Vaccine Refusal and Vaccine-Preventable Diseases in the United States: A Review of Measles and Pertussis [published correction appears in JAMA. 2016 May 17;315(19):2125] [published correction appears in JAMA. 2016 May 17;315 (19):2125]. JAMA. 2016;315(11):1149–1158. doi:10.1001/jama.2016.1353

7. Fisher KA, Bloomstone SJ, Walder J, Crawford S, Fouayzi H, Mazor KM. Attitudes Toward a Potential SARS-CoV-2 Vaccine: A Survey of U.S. Adults [published online ahead of print, 2020 Sep 4]. Ann Intern Med. 2020;M20–3569. doi:10.7326/M20-3569

8. Wheeler M, Buttenheim AM. Parental vaccine concerns, information source, and choice of alternative immunization schedules. Hum Vaccin Immunother. 2013;9(8):1782–1789. doi:10.4161/hv.25959

9. Salmon DA, Moulton LH, Omer SB, DeHart MP, Stokley S, Halsey NA. Factors associated with refusal of childhood vaccines among parents of school-aged children: a case-control study. Arch Pediatr Adolesc Med. 2005;159(5):470–476. doi:10.1001/archpedi.159.5.470

10. Omer SB, Salmon DA, Orenstein WA, deHart MP, Halsey N. Vaccine refusal, mandatory immunization, and the risks of vaccine-preventable diseases. N Engl J Med. 2009;360(19):1981–1988. doi:10.1056/NEJMsa0806477

11. Malik, A. A., Mcfadden, S. M., Elharake, J., & Omer, S. B. (2020). Determinants of COVID-19 vaccine acceptance in the US. EClinicalMedicine, 26, 100495. doi:10.1016/j.eclinm.2020.100495

12. Paterson P, Meurice F, Stanberry LR, Glismann S, Rosenthal SL, Larson HJ. Vaccine hesitancy and healthcare providers. Vaccine. 2016 Dec 20;34(52):6700–6.

13. Zhang J, While AE, Norman IJ. Nurses’ knowledge and risk perception towards seasonal influenza and vaccination and their vaccination behaviours: A cross-sectional survey. Int J Nurs Stud. 2011 Oct 1;48(10):1281–9.

14. LaVela SL, Smith B, Weaver FM, Legro MW, Goldstein B, Nichol K. Attitudes and Practices Regarding Influenza Vaccination Among Healthcare Workers Providing Services to Individuals With Spinal Cord Injuries and Disorders. Infect Control Hosp Epidemiol [Internet]. 2004 Nov [cited 2020 Oct 28];25(11):933–40. Available from: /core/journals/infection-control-and-hospital-epidemiology/article/attitudes-and-practices-regarding-influenza-vaccination-among-healthcare-workers-providing-services-to-individuals-with-spinal-cord-injuries-and-disorders/C672BE66C1D0649B3E0FA423C040AC91

15. Reiter PL, Pennell ML, Katz ML. Acceptability of a COVID-19 vaccine among adults in the United States: How many people would get vaccinated? Vaccine. 2020 Sep 29;38(42):6500–7.

16. Pogue K, Jensen JL, Stancil CK, Ferguson DG, Hughes SJ, Mello EJ, et al. Influences on Attitudes Regarding Potential COVID-19 Vaccination in the United States. Vaccines [Internet]. 2020 Oct 3 [cited 2020 Dec 15];8(4):582. Available from: https://www.mdpi.com/2076-393X/8/4/582

17. Fontanet A, Cauchemez S. COVID-19 herd immunity: where are we? [Internet]. Vol. 20, Nature Reviews Immunology. Nature Research; 2020 [cited 2020 Dec 18]. p. 583–4. Available from: https://www.defense.gouv.fr/content/download/583466/9938746/

18. Webb Hooper M, Nápoles AM, Pérez-Stable EJ. COVID-19 and Racial/Ethnic Disparities [Internet]. Vol. 323, JAMA - Journal of the American Medical Association. American Medical Association; 2020 [cited 2020 Dec 24]. p. 2466–7. Available from: https://www.chicago.gov/city/en/sites/covid-

19. Lemaitre M, Meret T, Rothan-Tondeur M, Belmin J, Lejonc JL, Luquel L, et al. Effect of influenza vaccination of nursing home staff on mortality of residents: A cluster-randomized trial. J Am Geriatr Soc [Internet]. 2009 Sep [cited 2020 Dec 24];57(9):1580–6. Available from: https://pubmed-ncbi-nlm-nih-gov.libproxy.unm.edu/19682118/

20. Zaffina S, Gilardi F, Rizzo C, Sannino S, Brugaletta R, Santoro A, et al. Seasonal influenza vaccination and absenteeism in health-care workers in two subsequent influenza seasons (2016/17 and 2017/18) in an Italian pediatric hospital. Expert Rev Vaccines [Internet]. 2019 Apr 3 [cited 2020 Dec 24];18(4):411–8. Available from: https://www-tandfonline-com.libproxy.unm.edu/doi/abs/10.1080/14760584.2019.1586541

21. Murti M, Otterstatter M, Orth A, Balshaw R, Halani K, Brown PD, et al. Measuring the impact of influenza vaccination on healthcare worker absenteeism in the context of a province-wide mandatory vaccinate-or-mask policy. Vaccine. 2019 Jul 9;37(30):4001–7.

22. Imai C, Toizumi M, Hall L, Lambert S, Halton K, Merollini K. A systematic review and meta-analysis of the direct epidemiological and economic effects of seasonal influenza vaccination on healthcare workers. Ortiz JR, editor. PLoS One [Internet]. 2018 Jun 7 [cited 2020 Dec 24];13(6):e0198685. Available from: https://dx.plos.org/10.1371/journal.pone.0198685

23. Romer D, Jamieson KH. Conspiracy theories as barriers to controlling the spread of COVID-19 in the U.S. Soc Sci Med. 2020 Oct 1;263:113356.

