## Supplemental Table 3 and Table 4 for "COVID-19 Vaccine Acceptance Among Health Care Workers in the United States"

Table 3: Likert scale – Supplemental Material

| | |
|---|---|
| **I would not get the vaccine because I do not believe they work** | |
| Strongly disagree | 2,493 (72%) |
| Disagree | 85 (2.4%) |
| Indifferent | 630 (18%) |
| Agree | 190 (5.5%) |
| Strongly agree | 81 (2.3%) |
| **I would not get the vaccine because I believe they are not safe** | |
| Strongly disagree | 2,120 (61%) |
| Disagree | 140 (4.0%) |
| Indifferent | 889 (26%) |
| Agree | 251 (7.2%) |
| Strongly agree | 79 (2.3%) |
| **I would not get the vaccine due to religious reasons** | |
| Strongly disagree | 2,974 (85%) |
| Disagree | 28 (0.8%) |
| Indifferent | 315 (9.1%) |
| Agree | 129 (3.7%) |
| Strongly agree | 33 (0.9%) |
| **I would not get the vaccine as it's my personal choice/freedom** | |
| Strongly disagree | 2,629 (76%) |
| Disagree | 187 (5.4%) |
| Indifferent | 398 (11%) |
| Agree | 171 (4.9%) |
| Strongly agree | 94 (2.7%) |
| **I would not get the vaccine as I fear needles** | |
| Strongly disagree | 3,001 (86%) |
| Disagree | 23 (0.7%) |
| Indifferent | 372 (11%) |
| Agree | 70 (2.0%) |
| Strongly agree | 13 (0.4%) |
| **I would not get the vaccine as I am worried about the effects of the COVID-19 Vaccine** | |
| Strongly disagree | 293 (8.4%) |
| Disagree | 344 (9.9%) |
| Indifferent | 427 (12%) |
| Agree | 1,441 (41%) |
| Strongly agree | 974 (28%) |
| **I would not get the vaccine as I am worried about the effectiveness of the Vaccine** | |
| Strongly disagree | 215 (6.2%) |

| | |
|---|---|
| Disagree | 324 (9.3%) |
| Indifferent | 540 (16%) |
| Agree | 1,607 (46%) |
| Strongly agree | 793 (23%) |
| **I would not get the vaccine as I am worried about the out of pocket cost coverage of the vaccine** | |
| Strongly disagree | 482 (14%) |
| Disagree | 734 (21%) |
| Indifferent | 1,340 (39%) |
| Agree | 678 (19%) |
| Strongly agree | 245 (7.0%) |
| **I would not get the vaccine due to the adverse effects of the vaccine on preexisting conditions** | |
| Strongly disagree | 1,511 (43%) |
| Disagree | 682 (20%) |
| Indifferent | 596 (17%) |
| Agree | 437 (13%) |
| Strongly agree | 253 (7.3%) |
| **I would not get the vaccine as there is no need for the vaccine for my risk level** | |
| Strongly disagree | 1,823 (52%) |
| Disagree | 939 (27%) |
| Indifferent | 452 (13%) |
| Agree | 167 (4.8%) |
| Strongly agree | 98 (2.8%) |
| **I would not get the vaccine as I have had prior reactions to other vaccinations** | |
| Strongly disagree | 2,316 (67%) |
| Disagree | 736 (21%) |
| Indifferent | 226 (6.5%) |
| Agree | 146 (4.2%) |
| Strongly agree | 55 (1.6%) |
| **I would not get the vaccine as I am worried about the rapidity of the COVID vaccine** | |
| Strongly disagree | 316 (9.1%) |
| Disagree | 288 (8.3%) |
| Indifferent | 319 (9.2%) |
| Agree | 1,238 (36%) |
| Strongly agree | 1,318 (38%) |
| **I would not get the vaccine as I do not trust pharmaceutical companies** | |
| Strongly disagree | 617 (18%) |
| Disagree | 861 (25%) |
| Indifferent | 964 (28%) |
| Agree | 658 (19%) |
| Strongly agree | 379 (11%) |

| | |
|---|---|
| **I would not get the vaccine as I do not trust the FDA/CDC** | |
| Strongly disagree | 586 (17%) |
| Disagree | 727 (21%) |
| Indifferent | 631 (18%) |
| Agree | 903 (26%) |
| Strongly agree | 632 (18%) |
| **I would not get the vaccine as I do not trust the doctors recommending the vaccine** | |
| Strongly disagree | 1,481 (43%) |
| Disagree | 1,046 (30%) |
| Indifferent | 650 (19%) |
| Agree | 201 (5.8%) |
| Strongly agree | 101 (2.9%) |
| **I would not get the vaccine as I do not trust the government about the COVID-19 severity** | |
| Strongly disagree | 666 (19%) |
| Disagree | 594 (17%) |
| Indifferent | 621 (18%) |
| Agree | 792 (23%) |
| Strongly agree | 806 (23%) |

Table 4: Likert scale – breakdown Supplemental Material

| | **No**, N = 279[1] | **Wait for Review**, N = 1,953[1] | **Yes**, N = 1,247[1] | **p-value**[2] |
|---|---|---|---|---|
| **I would not get the vaccine because I do not believe they work** | | | | <0.001* |
| Strongly disagree | 81 (29%) | 1,351 (69%) | 1,061 (85%) | |
| Disagree | 86 (31%) | 425 (22%) | 119 (9.5%) | |
| Indifferent | 71 (25%) | 99 (5.1%) | 20 (1.6%) | |
| Agree | 23 (8.2%) | 45 (2.3%) | 17 (1.4%) | |
| Strongly agree | 18 (6.5%) | 33 (1.7%) | 30 (2.4%) | |
| **I would not get the vaccine because I believe they are not safe** | | | | <0.001* |
| Strongly disagree | 61 (22%) | 1,078 (55%) | 981 (79%) | |
| Disagree | 80 (29%) | 610 (31%) | 199 (16%) | |
| Indifferent | 63 (23%) | 160 (8.2%) | 28 (2.2%) | |
| Agree | 40 (14%) | 82 (4.2%) | 18 (1.4%) | |
| Strongly agree | 35 (13%) | 23 (1.2%) | 21 (1.7%) | |
| **I would not get the vaccine due to religious reasons** | | | | 0.161 |
| Strongly disagree | 139 (50%) | 1,668 (85%) | 1,167 (94%) | |
| Disagree | 62 (22%) | 204 (10%) | 49 (3.9%) | |
| Indifferent | 53 (19%) | 56 (2.9%) | 20 (1.6%) | |
| Agree | 13 (4.7%) | 9 (0.5%) | 6 (0.5%) | |
| Strongly agree | 12 (4.3%) | 16 (0.8%) | 5 (0.4%) | |
| **I would not get the vaccine as it's my personal choice/freedom** | | | | <0.001* |
| Strongly disagree | 63 (23%) | 1,452 (74%) | 1,114 (89%) | |
| Disagree | 37 (13%) | 280 (14%) | 81 (6.5%) | |
| Indifferent | 45 (16%) | 98 (5.0%) | 28 (2.2%) | |
| Agree | 71 (25%) | 100 (5.1%) | 16 (1.3%) | |
| Strongly agree | 63 (23%) | 23 (1.2%) | 8 (0.6%) | |
| **I would not get the vaccine as I fear needles** | | | | 0.749 |
| Strongly disagree | 185 (66%) | 1,654 (85%) | 1,162 (93%) | |
| Disagree | 60 (22%) | 243 (12%) | 69 (5.5%) | |

| | | | | |
|---|---|---|---|---|
| Indifferent | 24 (8.6%) | 35 (1.8%) | 11 (0.9%) | |
| Agree | 8 (2.9%) | 14 (0.7%) | 1 (<0.1%) | |
| Strongly agree | 2 (0.7%) | 7 (0.4%) | 4 (0.3%) | |
| **I would not get the vaccine as I am worried about the effects of the COVID-19 Vaccine** | | | | <0.001* |
| Strongly disagree | 2 (0.7%) | 47 (2.4%) | 244 (20%) | |
| Disagree | 0 (0%) | 56 (2.9%) | 288 (23%) | |
| Indifferent | 13 (4.7%) | 175 (9.0%) | 239 (19%) | |
| Agree | 40 (14%) | 980 (50%) | 421 (34%) | |
| Strongly agree | 224 (80%) | 695 (36%) | 55 (4.4%) | |
| **I would not get the vaccine as I am worried about the effectiveness of the Vaccine** | | | | 0.11 |
| Strongly disagree | 2 (0.7%) | 49 (2.5%) | 164 (13%) | |
| Disagree | 3 (1.1%) | 103 (5.3%) | 218 (17%) | |
| Indifferent | 31 (11%) | 283 (14%) | 226 (18%) | |
| Agree | 58 (21%) | 996 (51%) | 553 (44%) | |
| Strongly agree | 185 (66%) | 522 (27%) | 86 (6.9%) | |
| **I would not get the vaccine as I am worried about the out of pocket cost coverage of the vaccine** | | | | <0.001* |
| Strongly disagree | 112 (40%) | 617 (32%) | 611 (49%) | |
| Disagree | 42 (15%) | 418 (21%) | 274 (22%) | |
| Indifferent | 79 (28%) | 437 (22%) | 162 (13%) | |
| Agree | 16 (5.7%) | 318 (16%) | 148 (12%) | |
| Strongly agree | 30 (11%) | 163 (8.3%) | 52 (4.2%) | |
| **I would not get the vaccine due to the adverse effects of the vaccine on preexisting conditions** | | | | 0.155 |
| Strongly disagree | 71 (25%) | 707 (36%) | 733 (59%) | |
| Disagree | 44 (16%) | 387 (20%) | 251 (20%) | |
| Indifferent | 75 (27%) | 388 (20%) | 133 (11%) | |
| Agree | 34 (12%) | 297 (15%) | 106 (8.5%) | |
| Strongly agree | 55 (20%) | 174 (8.9%) | 24 (1.9%) | |
| **I would not get the vaccine as there is no need for the vaccine for my risk level** | | | | <0.001* |
| Strongly disagree | 24 (8.6%) | 904 (46%) | 895 (72%) | |
| Disagree | 37 (13%) | 649 (33%) | 253 (20%) | |
| Indifferent | 93 (33%) | 288 (15%) | 71 (5.7%) | |
| Agree | 66 (24%) | 81 (4.1%) | 20 (1.6%) | |
| Strongly agree | 59 (21%) | 31 (1.6%) | 8 (0.6%) | |

| | | | | |
|---|---|---|---|---|
| **I would not get the vaccine as I have had prior reactions to other vaccinations** | | | | 0.005* |
| Strongly disagree | 99 (35%) | 1,202 (62%) | 1,015 (81%) | |
| Disagree | 73 (26%) | 482 (25%) | 181 (15%) | |
| Indifferent | 58 (21%) | 140 (7.2%) | 28 (2.2%) | |
| Agree | 30 (11%) | 98 (5.0%) | 18 (1.4%) | |
| Strongly agree | 19 (6.8%) | 31 (1.6%) | 5 (0.4%) | |
| **I would not get the vaccine as I am worried about the rapidity of the COVID vaccine** | | | | <0.001* |
| Strongly disagree | 7 (2.5%) | 69 (3.5%) | 240 (19%) | |
| Disagree | 7 (2.5%) | 65 (3.3%) | 216 (17%) | |
| Indifferent | 24 (8.6%) | 143 (7.3%) | 152 (12%) | |
| Agree | 37 (13%) | 714 (37%) | 487 (39%) | |
| Strongly agree | 204 (73%) | 962 (49%) | 152 (12%) | |
| **I would not get the vaccine as I do not trust pharmaceutical companies** | | | | 0.029* |
| Strongly disagree | 5 (1.8%) | 185 (9.5%) | 427 (34%) | |
| Disagree | 15 (5.4%) | 424 (22%) | 422 (34%) | |
| Indifferent | 67 (24%) | 636 (33%) | 261 (21%) | |
| Agree | 66 (24%) | 480 (25%) | 112 (9.0%) | |
| Strongly agree | 126 (45%) | 228 (12%) | 25 (2.0%) | |
| **I would not get the vaccine as I do not trust the FDA/CDC** | | | | 0.019* |
| Strongly disagree | 3 (1.1%) | 161 (8.2%) | 422 (34%) | |
| Disagree | 9 (3.2%) | 367 (19%) | 351 (28%) | |
| Indifferent | 52 (19%) | 397 (20%) | 182 (15%) | |
| Agree | 67 (24%) | 609 (31%) | 227 (18%) | |
| Strongly agree | 148 (53%) | 419 (21%) | 65 (5.2%) | |
| **I would not get the vaccine as I do not trust the doctors recommending the vaccine** | | | | <0.001* |
| Strongly disagree | 22 (7.9%) | 604 (31%) | 855 (69%) | |
| Disagree | 46 (16%) | 695 (36%) | 305 (24%) | |
| Indifferent | 96 (34%) | 487 (25%) | 67 (5.4%) | |
| Agree | 53 (19%) | 132 (6.8%) | 16 (1.3%) | |
| Strongly agree | 62 (22%) | 35 (1.8%) | 4 (0.3%) | |
| **I would not get the vaccine as I do not trust the government about the COVID-19 severity** | | | | 0.59 |
| Strongly disagree | 7 (2.5%) | 255 (13%) | 404 (32%) | |
| Disagree | 16 (5.7%) | 334 (17%) | 244 (20%) | |

| | | | |
|---|---|---|---|
| Indifferent | 55 (20%) | 374 (19%) | 192 (15%) |
| Agree | 55 (20%) | 503 (26%) | 234 (19%) |
| Strongly agree | 146 (52%) | 487 (25%) | 173 (14%) |
